# Impaired Scaling of Step Length in Parkinsonian Postural Instability

**DOI:** 10.1101/2020.08.25.20179945

**Authors:** Robert A. McGovern, Juan C. Cortés, Anne P. Wilson, Guy M. McKhann, Pietro Mazzoni

## Abstract

**Background:** Postural stepping is an important strategy for recovery of balance in response to postural perturbations. It is disrupted by Parkinson’s disease (PD) and other conditions. The nature of this disruption remains poorly understood. Understanding the motor control nature of this impairment can guide the development of novel interventions.

**Objectives:** To identify the motor control abnormalities responsible for parkinsonian impairment of postural stepping.

**Methods:** We studied four groups of participants: control, aged, PD, and normal-pressure hydrocephalus (NPH). We performed kinematic analysis of postural stepping by recording participants’ body motion during a modified version of the clinical pull test, which was performed multiple times with different amounts of pulling forcefulness.

**Results:** Successful postural stepping in the control group was accompanied by linear scaling of their first step’s length and latency to the body’s initial motion: more forceful pulls caused larger initial body acceleration, which resulted in longer steps that began earlier. PD patients exhibited reduced scaling of step length: they maintained normal reaction time but took steps that were inadequately short. Reduced step length scaling was present, but less severe, in aged individuals, and was more severe in NPH patients. Aged individuals and PD patients exhibited partial compensation for reduced step length scaling: their step length included a component that was independent of initial body acceleration, which was absent in control and NPH groups.

**Conclusions:** the impairment of postural stepping caused by PD and related conditions is due to inadequate scaling of movement amplitude and is thus a form of hypokinesia.

## Introduction

When standing in a bus that starts moving, we must sometimes take a step back to avoid falling. This behavior is an example of the postural stepping response, an important motor strategy that helps us maintain balance in response to postural perturbations.

We can counter small perturbations by adjusting our body’s geometry without moving our feet (hip and ankle strategies) [1]. These strategies are inadequate, however, for a variety of perturbations encountered in daily life, such as stumbling on a curb, being nudged when standing in a crowd, and stepping off an escalator. Postural stepping is a commonly used strategy when there are no constraints on foot movement [2]. It can counter a wide variety of perturbation types and magnitudes [3,4], as it allows balance recovery by changing the body’s base of support.

Balance disorders are an important cause of falls, which in turn are a major cause of morbidity, mortality, and reduced quality of life in the elderly and in patients with neurologic disorders [5–7]. The balance disorder caused by Parkinson’s disease (PD) and related conditions (parkinsonism) is characterized by impaired postural stepping, as exhibited in inadequate responses to the pull test [8]. The nature of this impairment, however, remains unclear.

Studies of postural stepping in PD patients [9–14] have identified reduced static sway before stepping [9], particular susceptibility to backwards perturbations [15], and abnormally coordinated responses (simultaneous activation of ankle, hip and trunk muscles) to surface force plate translation [9,11,12]. PD patients also exhibit anticipatory lateral postural adjustments, later-onset and shorter steps [16,17], increased weight shift time, and a base-width neutral step [18]. These varied abnormalities do not readily point to a motor control problem responsible for impaired postural stepping.

In spite of the complex sensory processing and precisely coordinated force control required to maintain balance, postural stepping can be described rather simply at the task level in kinematic terms: as the body moves away from its base of support, a step is taken to extend the base of support and counter the body’s falling motion. The relevant perturbation is the body’s movement away from a stable configuration, regardless of the nature and magnitude of the force that caused this perturbation. The relevant response is placement of the foot in the appropriate position and at the appropriate time to counter the body’s motion. Successful postural stepping may thus be explained by a kinematic account, i.e. by relationships between initial body motion and kinematic features of the stepping response.

An interesting feature of normal postural stepping is that, when pulled backwards at the waist with different force magnitudes, the step length of control participants increases as force increases [19]. This finding suggests that a scaling relationship exists between magnitude of postural perturbation and the stepping response. We thus hypothesized that successful postural stepping can be described by a kinematic control policy: when posture is perturbed, a step is taken with appropriate amplitude and latency to stop the body’s motion.

If normal postural stepping can be described kinematically, then perhaps its impairment seen in PD and other conditions has a kinematic explanation: are the steps of postural stepping too short, too slow, or too late, to stop the body from falling? Such a kinematic account of postural stepping impairment would link parkinsonian postural instability to bradykinesia, a complex of motor symptoms that includes slowness (bradykinesia itself), reduced amplitude (hypokinesia), and delay in movement onset (akinesia) [20].

We devised a quantitative version of a clinical maneuver commonly used to test the stepping response, the pull test, in which a participant is pulled from behind and must step backwards to recover balance [8]. By recording the body’s motion in response to repeated pulls of varying intensity, we examined relationships between postural perturbation intensity and stepping kinematics. We characterized the kinematic control policy of normal postural stepping in control participants, and then examined how this control is disrupted by PD and by two other conditions that disrupt postural responses, normal aging and normal pressure hydrocephalus (NPH).

## Methods

### Participants

We studied 29 participants with no musculoskeletal disorder, dementia, or depression (Table 1): control participants (*Control* group); older participants (*Aged* group); patients with PD based on UK Brain Bank Criteria [21] (*PD* group); patients with NPH (inclusion criteria: progressive gait impairment with multiple falls, cognitive symptoms, urinary incontinence, and communicating hydrocephalus disproportionate to cerebral atrophy). PD patients were tested on their usual medications. The study protocol was approved by the Institutional Review Board.

### Quantitative Pull Test Protocol

Participants were pulled from behind by one of us (P.M.) with a firm brisk pull at the shoulders [22] and were caught if they did not recover balance on their own. This procedure was repeated for 8-20 trials with varying degrees of pull intensity. A Proreflex camera (Qualisys AB) captured motion of the right shoulder, elbow, hip, knee, toe, and heel in the sagittal plane. Participants held their arms folded to prevent reflective marker occlusion.

### Data Analysis

We recorded the number of steps (steps taken before stopping) and failure rate (fraction of trials without recovery after 3 steps). We calculated the body’s center of mass (*COM*) position using standard biomechanical equations [23]. We differentiated COM and foot position to obtain COM and foot velocity and acceleration.

A pull test trial consisted of an initial backward motion of the upper body (*perturbation phase*; Fig. 1A), followed by one or more backward steps (*response phase*; Fig 1B). Pull onset was the time when COM acceleration exceeded 15 cm/s^2^. Step onset and landing were the times when the ankle marker’s velocity crossed a 3 cm/s threshold for the first step (Fig. 1C). We quantified perturbation intensity as the average COM acceleration after pull onset and before step onset (shaded area in lower panel of Fig. C). The response (first step) was characterized by reaction time (time to step onset; Fig. 1C), horizontal amplitude (step length), and duration (time from step onset to step landing).

**Figure 1.**
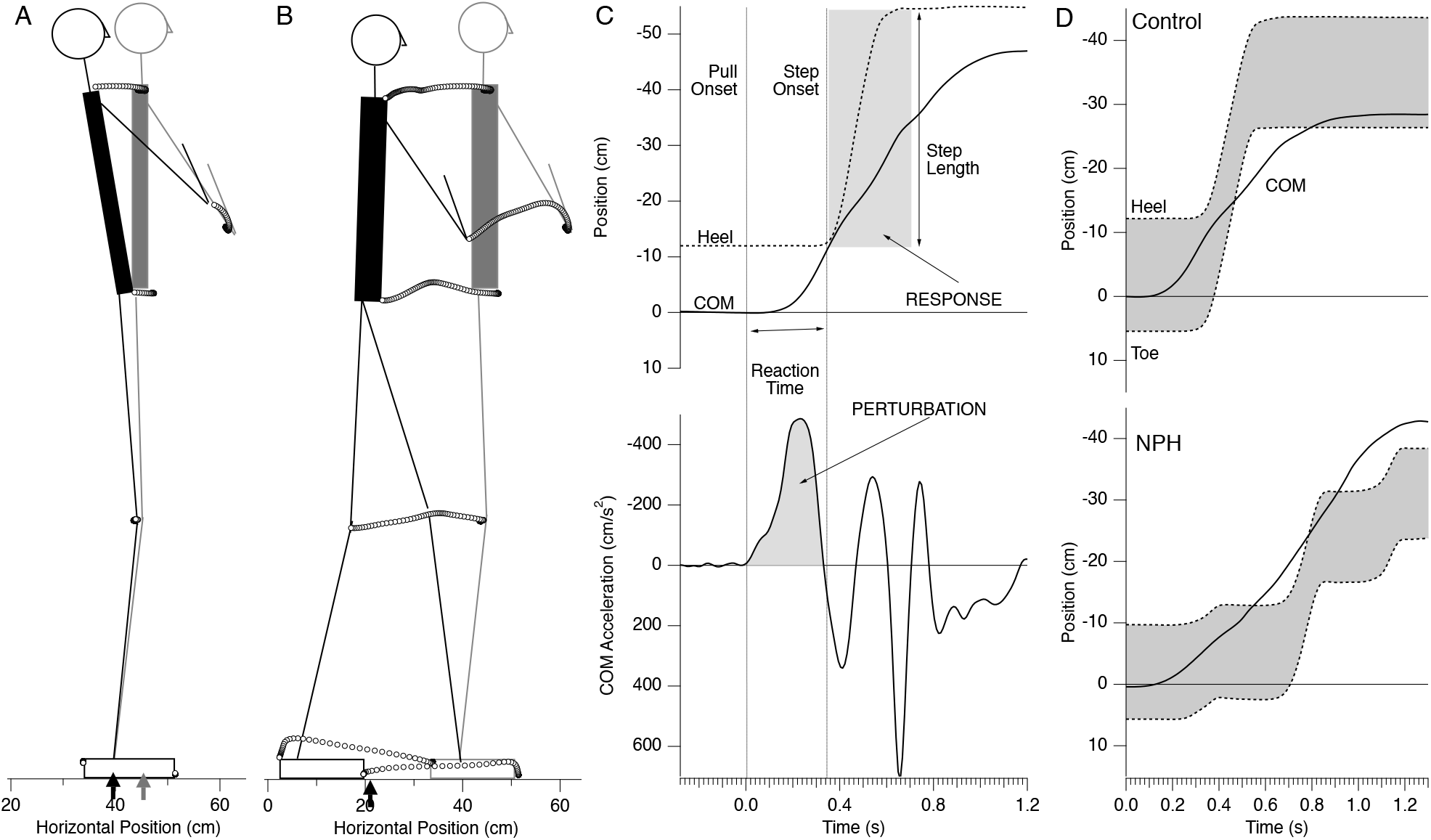
Trajectories of motion capture markers from pull onset (t=0; grey) to t=250 ms (black) in a pull test trial. Arrows indicate horizontal center-of-mass (COM) position. **B**. Marker trajectories up to time of first step landing (t=670 ms). **C**. Kinematic quantities in a pull test trial for a CTL participant. **D**. Horizontal position of COM (solid), heel, toe (dashed) during a single trial in individual CTL (upper traces) and NPH (lower traces) participants.

### Statistical Analysis

We computed linear correlation between selected kinematic variables. We performed ANOVA with corrections for multiple comparisons using Tukey’s method of Honest Significant Differences (JMP, SAS Institute), and with significance set at alpha=0.05.

## Results

Participants did not differ in sex ratio, height, weight, or cognitive scores (Montreal Cognitive Assessment [24]). Control participants were younger than other groups, as intended (Table 1). PD and NPH participants had similar UPDRS III scores. Pull test scores differed across groups (ANOVA, p<0.0001): Control and Aged participants exhibited normal recovery (pull test score 0); PD and NPH had higher pull test scores than Control and Aged groups (*t* test, p<0.05); and NPH had higher pull test scores than PD participants (*t* test, p=0.002; Table 1).

Participants differed in number of steps (steps taken before stopping) and failure rate (fraction of trials without recovery after 3 steps; Table 1; ANOVA, p<0.001). Control participants all recovered in one step in all trials, regardless of pull intensity. Aged and PD groups more steps than Control (p<0.05) and less than NPH (p<0.01). NPH participants failed more frequently than Control and Aged p<0.01).

Because all control participants recovered their balance in once step, we focused our analysis on the first step. We looked for group differences in first step features that might explain why participants in PD and NPH groups had higher failure rates.

The kinematics of a successful trial illustrate how postural stability may be regained after a pull. The COM initially accelerated backward after a pull, away from its initial position under the base of support (arrows in Fig. 1A; upper panel of Fig. 1D). The first step landed well behind the COM, which stopped the COM’s backward motion and restored postural stability (Fig. 1B; upper panel of Fig. 1D). Failure to recover is illustrated in a trial for an NPH participant (Fig. 1D, lower panel). After a short first step, the COM continued to move backwards in spite of additional short steps.

Initial COM acceleration values varied widely, as intended, within each participant, and ranged between 20 and 340 cm/s^2^. We examined the time course of COM velocity after the first step to establish how quickly participants regained their balance in a subset of trials with matched initial COM acceleration. Control participants’ speed decreased from the end of the first step (t=0) to near 0 at t=1 s (Fig. 2A). Aged participants’ speed decreased to the same value as Control at t=1 s (Tukey HSD; p>0.1), though their intermediate speeds were higher than for Control (p=0.0003 at 0.25s; p<0.0001 at 0.5s). PD participants’ speed was higher than for Control (p=0.0002) and Aged (p<0.05) groups at t=1 s. NPH participants’ speed remained the highest across groups at t=1 s (p<0.0001 vs PD group speed). Thus, PD and NPH participants were still moving 1 second after the first step ended. These results are consistent with the groups’ rankings in number of steps and failure rates (Table 1).

**Figure 2.**
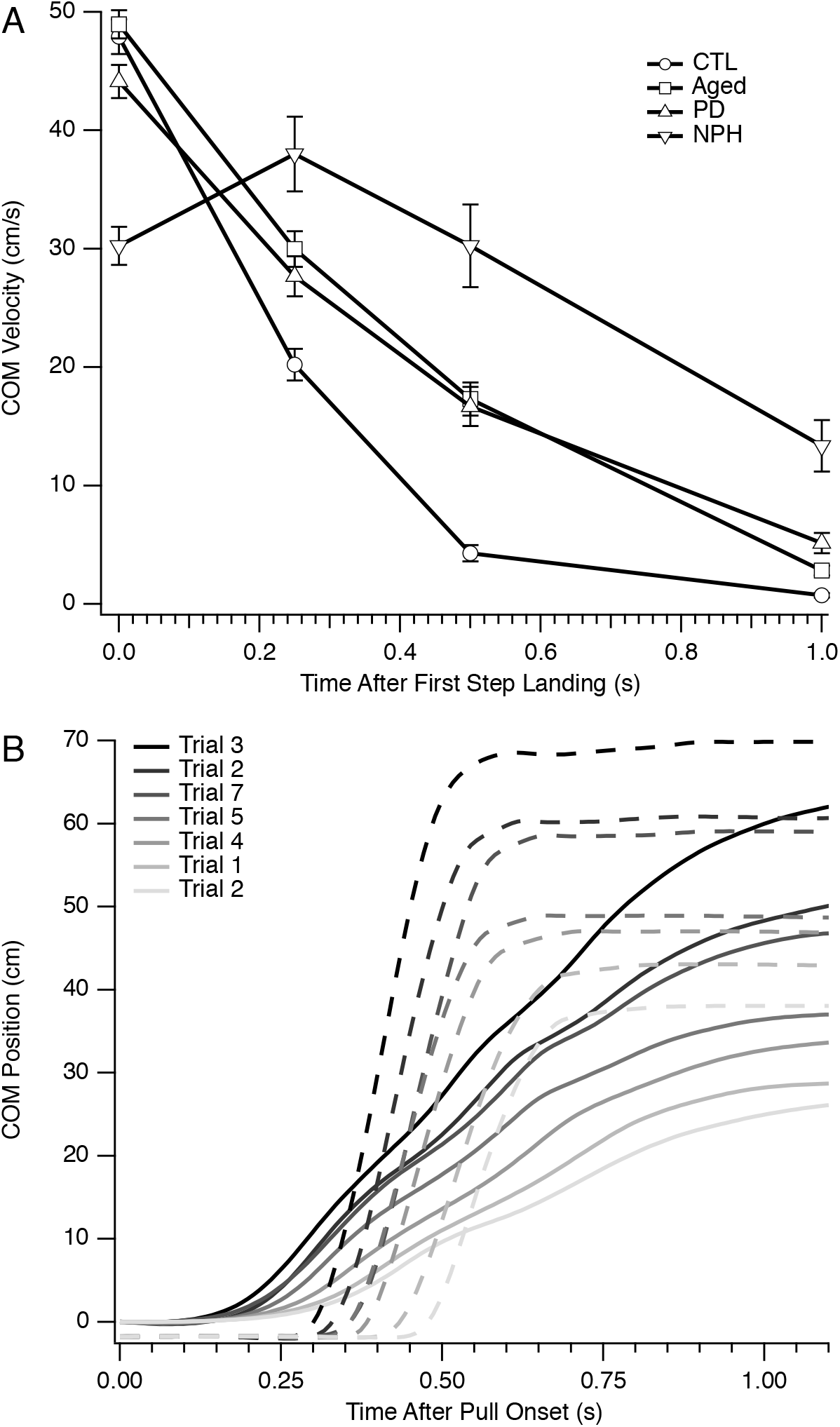
**A**. COM velocity after first step landing for trials with average COM acceleration matched across groups. **B**. Time course of COM and foot (heel) position for selected trials of different perturbation magnitude (randomly interleaved) in a CTL participant.

Pulls of greater intensity caused faster and larger changes in COM position (Fig. 2B, upper panel), which were followed by steps of earlier onset, shorter duration, and larger amplitude (Fig. 2B, lower panel). This finding suggests a kinematic control policy for postural stepping: step latency and amplitude are scaled to the intensity of postural perturbation, so that the foot lands sufficiently behind the COM to stop its motion. We examined Control participants’ responses for evidence of this policy and then examined how it might be disrupted in the other groups.

There was a marked linear correlation between step length and initial COM acceleration, both within and across Control participants (Figure 3A, 3B; *r* = 0.80 ± 0.2, mean ±SEM; p<0.01 for each participant). Similarly, there was an inverse correlation between reaction time and initial COM acceleration, (Figure 4A, 4B; *r* = 0.68 ± 0.2; p<0.05 in each participant). By contrast, first step duration did not significantly vary across trials (ANOVA, p>0.05, mean *r* = 0.35 + 0.2). These correlations are a plausible mechanism for stepping responses’ success: larger perturbations are handled by making the first step of larger amplitude and earlier onset, so that the foot lands further behind the moving body and has a better chance of halting its motion.

**Figure 3.**
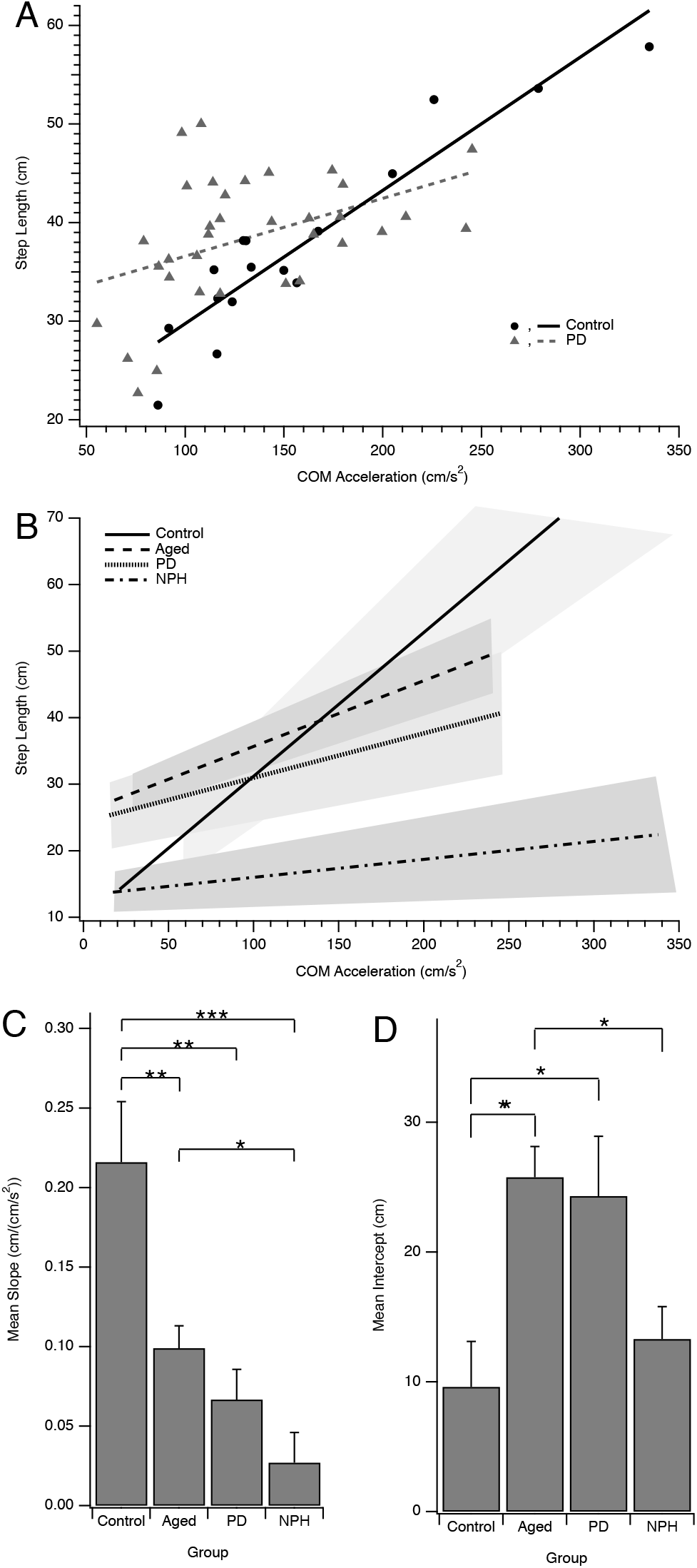
**A**. Step length vs. initial COM acceleration for all trials for one CTL and one PD participant. Lines = linear correlation. **B**. Correlation lines between step length and initial COM acceleration. Shading = standard error. **C**. Slope (mean ± SEM) of linear correlation between step length and initial COM acceleration. *, p<0.05; **, p<0.01; ***, p<0.001 (Tukey HSD contrasts). **D**. Intercept (mean ± SEM) of linear regression between step length and initial COM acceleration.

**Figure 4.**
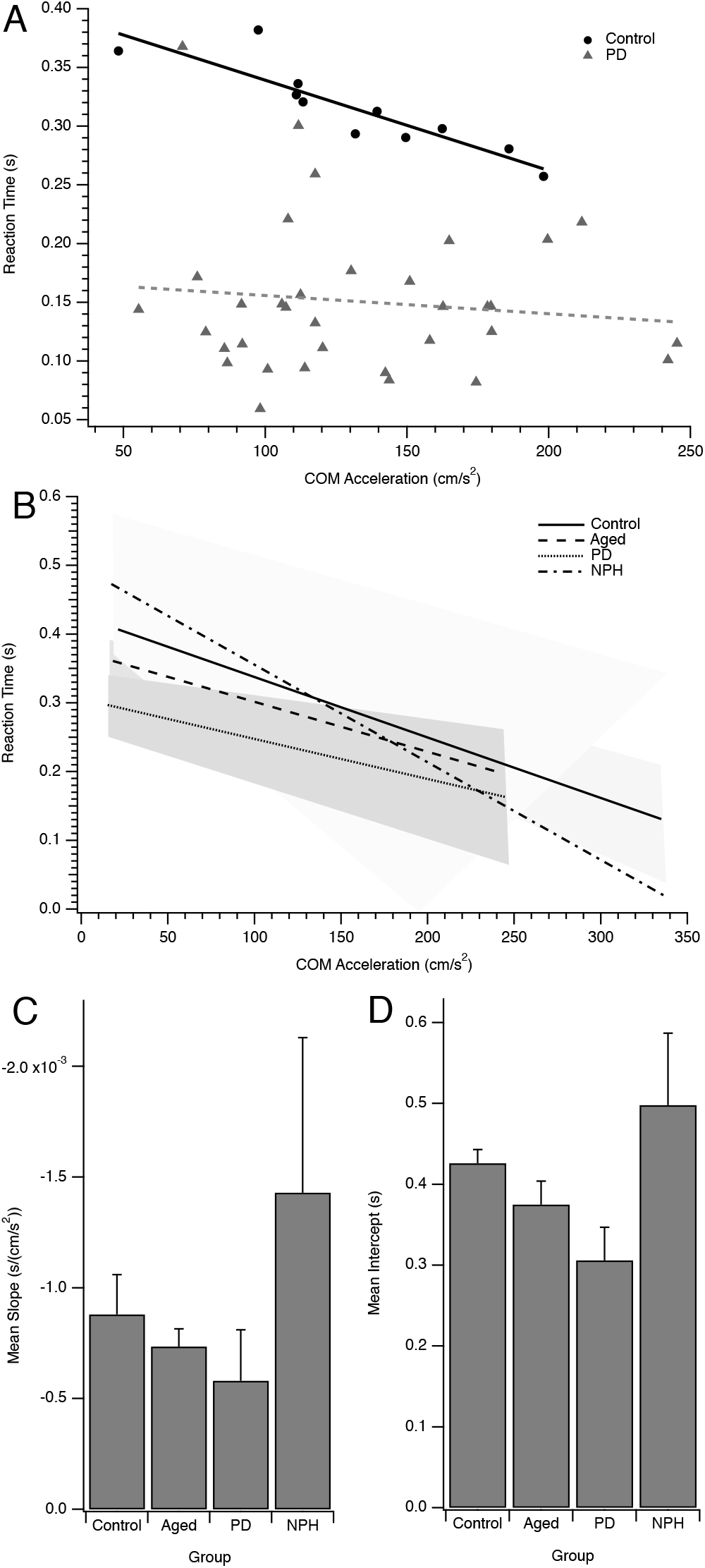
**A**. Reaction time vs. initial COM acceleration for all trials for one CTL and one PD participant. Lines = linear correlation. **B**. Correlation lines between reaction time and initial COM acceleration for each participant group. **C**. Slope (mean ± SEM) of linear regression between reaction time and initial COM acceleration. *, p<0.05; **, p<0.01; ***, p<0.001 (Tukey HSD contrasts). **D**. Intercept (mean ± SEM) of linear regression between reaction time and initial COM acceleration.

The correlation of step length and initial COM acceleration is characterized by a slope and a y-intercept. The slope indicates how much longer the step is in response a larger initial acceleration and represents a scaling factor between acceleration and step length. The correlation between step length and initial COM acceleration had a lower slope in the Aged, PD, and NPH groups compared to Control (Figure 3B, 3C; ANOVA, p<0.0001, Control vs. Aged, Tukey HSD, p=0.002, Control vs. PD, Tukey HSD, p=0.0003, Control vs. NPH, Tukey HSD, p<0.0001). Aged participants had a higher slope than NPH participants (Aged vs. NPH, Tukey HSD, p=0.04), but otherwise slope was not significantly different among Aged, PD, and NPH groups (p>0.05 for all other comparisons, Tukey HSD).

The correlation’s y-intercept indicates, in practice, the smallest possible step that a participant takes in response to the smallest possible initial COM acceleration. It raises the entire correlation line by this amount, and thus indicates a bias--a minimum amount of step length that is added to every step, regardless of perturbation magnitude. The step length correlation’s intercept was higher in Aged and PD groups compared to Control (Figure 3B, 3D; Aged vs. Control, Tukey HSD, p=0.006, PD vs. Control, Tukey HSD, p=0.02), and smaller in the NPH group compared to Aged (Tukey HSD, p=0.04).

The lower slope values indicate that the step length of Aged, PD, and NPH participants did not increase by a normal amount in response to increasing perturbations. For sufficiently large COM accelerations, step length of Aged and PD participants was smaller than for Control (Figure 4B). Step length in the NPH group was shorter than for the Control group for most of the range of COM accelerations (Figure 4B). The higher intercept values of Aged and PD participants, on the other hand, made their steps longer than they would have otherwise been. As a result, for Aged and PD groups, the first step was longer than necessary in the range of smaller COM accelerations, and less abnormally short for larger COM accelerations (Figure 4B). An increase in the intercept thus countered the effect of slope reduction and partially compensated for the loss of adequate scaling of step size to perturbation magnitude in Aged and PD groups. For example, step length for the PD group was shorter than normal only for accelerations above ~100 cm/s^2^ (Figure 4B) because of the higher intercept of this group’s correlation. This effect was not seen in the NPH group.

The correlation between reaction time and initial COM acceleration did not significantly vary across groups (Figure 4; slope, ANOVA, p=0.45, all Tukey HSD comparisons, p>0.45; intercept, ANOVA, p=0.10, all Tukey HSD comparisons, p>0.08).

## Discussion

Kinematic analysis of a clinical test of postural stability revealed that the normal response to postural perturbations across a wide range of intensities is to recover by scaling the first step’s amplitude and latency to the body’s initial acceleration. These scaling relationships indicate a *kinematic strategy* for successful recovery from postural perturbations of different intensities: increasing step length and decreasing reaction time allows the body to recover balance by placing the foot further behind the center of mass, and sooner, in response to greater postural perturbations.

Aged participants successfully recovered but took more than one step. PD and NPH participants took more steps and had higher failure rates. The slope of the correlation between step length and postural perturbation magnitude was reduced in all 3 groups, and its intercept was increased in Aged and PD groups. Reaction time scaling to postural perturbation was normal in Aged, PD, and NPH groups.

The slope reduction in step length scaling in Aged, PD, and NPH groups offers a kinematic explanation for failure to recover balance in the pull test. For larger perturbations, step size was abnormally small, so that the foot was not placed far enough behind the body to stop its motion.

Aged participants exhibited the same type of postural stepping abnormality, reduced step length scaling, as PD participants, though this reduction was less than in the PD group. Aged and PD groups also exhibited a compensatory increase in the intercept of the correlation between body acceleration and step size. This compensatory change was of similar magnitude in both groups. It may have been sufficient to counter the smaller amount of slope reduction in the Aged group and thus explain their lower failure rate in the pull test (Table 1). The intercept increase may have not been sufficient to counter the larger slope reduction of the PD group, which potentially accounts for the group’s higher failure rate.

Regarding the symptom complex of bradykinesia (bradykinesia proper, hypokinesia, akinesia; see Introduction) [20], our findings indicate a postural stepping abnormality that consists of hypokinesia: steps were shorter than they needed to be to allow balance recovery in PD and NPH groups. By contrast, these participants did not show evidence of delayed movement onset (akinesia). This finding could reflect normal control of step reaction time in the groups we studied, or a combined effect of reaction time increase, known to be caused by PD [25] and compensatory reaction time reduction to counteract first step hypokinesia. Similarly, there was no evidence of movement slowing.

Finding a similar motor control abnormality underlying abnormal postural stepping across Aged, PD, and NPH groups suggests that these conditions share brain changes responsible for postural instability. A shared pathology for PD and NPH is consistent with the overlap of other clinical manifestations of these disorders, such as bradykinesia of rapid repeated leg and foot movements [26]. Similarly, aging is accompanied by brain changes also seen in PD, including loss of neurons in the substantia nigra and parkinsonian movement abnormalities [27]. In aging, however, these abnormalities may be subclinical, rather than entirely absent, thanks to compensatory mechanisms like the rise in the intercept of the step length scaling relationship.

The lack of evidence of compensation (increase in intercept) in the NPH group may indicate that compensatory mechanisms are not available when kinematic scaling relationships are severely disrupted: NPH participants’ slopes for the step size vs. initial acceleration scaling were nearly flat. Alternatively, NPH may disrupt additional gait control mechanisms not affected by aging or PD.

Elderly individuals are known to take multiple steps when recovering from a postural perturbation [28–31]. Step kinematics differed from those of young participants in some studies [29,31] but were normal in another study [30]. Our results may explain this variation as emerging from the changes in the scaling relationship of step length to perturbation magnitude. Thanks to the compensatory effect of increased intercept, aged participants in our study took abnormally short steps only for perturbations greater than 150 cm/s^2^ (Fig. 3B).

PD patients have been reported to take abnormally short steps in compensatory stepping, as we found, but also to exhibit increased reaction time, unlike our findings [17]. Differences in how perturbations were applied may account for this discrepancy: platform motion is likely more sudden than a manual pull at the shoulders, and could thus unmask reaction time increases not visible in the pull test. Another study [18] did not find an effect of PD on step length, which we also found for initial body accelerations below 100 cm/s^2^. Some of these findings, such as reduced static sway, may reflect the same kinematic abnormality (hypokinesia) we identified, while others may be independent additional contributors to impaired postural stepping.

Postural response impairment in PD has been examined within a model of standing as the balancing of a two-segment inverted pendulum, in which the CNS acting as a multivariate feedback controller that processes sensory information, estimates body kinematics, and sends appropriate motor commands scaled by feedback gains [32]. PD patients in this study had abnormal feedback gains and were unable to scale postural responses to changes in perturbation amplitude. Although these results were observed in a fixed-support postural task, the model offers a convincing mechanism to explain our findings for normal and abnormal stepping responses. Consistent with this model, our findings show that the normal stepping response is governed by a linear gain between perturbation magnitude and postural response, and that PD reduces this gain so that postural responses are scaled down.

While gait parameters have been studied in detail for NPH patients [33], our understanding of postural control in these patients remains limited. NPH patients appear to have a larger static sway area and higher backwards directed COM velocity during upright stance [34], but no dynamic studies have examined NPH patients’ postural control in detail. The patients in our study were essentially unable to scale step length to increases in perturbation amplitude. This resulted in the highest percentage of inadequate responses among the groups we studied.

We chose the COM’s initial backward acceleration as the measure of perturbation intensity, rather than the force applied at the shoulders. The COM’s initial acceleration results from the combined effect of pull force and inertial resistance due to a person’s mass, height, posture, and stiffness (mechanical impedance). COM acceleration thus reflects the *effective* intensity of the pull, that is, how effectively a pull’s force displaces the body away from a stable posture. We would argue that the relevant variable that ultimately leads to loss of balance is the body’s motion towards the limits of the base of support, and not the force applied at the shoulders. The body’s motion is what needs to be countered: a fall is, after all, the end-result of the COM being in the wrong position, and headed in the wrong direction, relative to the base of support for too long. Therefore, even though COM acceleration is not an independent variable (as it is an outcome of the applied force), we consider it an appropriate measure of perturbation intensity in the pull test.

An advantage of using COM acceleration as a measure of perturbation intensity is that it is not confounded by anticipatory strategies. If participants leaned forward or stiffened their body in anticipation of the pull, the effect of a given pull force would be diminished. These strategies are equivalent to increasing inertial resistance. Their dampening effect on applied force is thus accurately reflected in the COM’s initial acceleration and does not confound the estimate of perturbation intensity.

A kinematic explanation of postural instability as a manifestation of hypokinesia suggests that postural responses should improve with treatments that benefit other forms of hypokinesia. At this time, whether any treatment improves postural instability is in our opinion unclear. Treatment of PD with levodopa is associated with reduced frequency of falls [35] and with improved scores on all motor subcomponents of the UPDRS, including the subscale related to postural control and gait [36]. However, levodopa worsened the ability to scale responses to large perturbation amplitudes in a fixed-support strategy postural task [37]. Postural instability has been reported not to benefit from deep brain stimulation (DBS) when assessed with clinical measures [38], but showed clear benefit from DBS (Nantel *et al*., 2012) and pallidotomy [41] when assessed with posturography. Compensatory stepping has been shown to be unaffected by globus pallidus internus DBS while subthalamic nucleus DBS has been associated with delays in the preparatory phase prior to stepping and more steps required to regain balance [42]. Treatment of NPH with ventriculoperitoneal shunting has benefit on gait [43,44]; whether this treatment improves postural responses is unclear.

We found that successful recovery of balance in postural stepping is mediated by a kinematic mechanism (scaling of first step length and reaction time to initial body motion) and that parkinsonian postural instability can be explained by hypokinesia of the first step. People with parkinsonism have difficulty recovering balance because their reactive steps are too small relative to the size of the imposed perturbation, and not because of a delay in step initiation. Further quantitative testing of postural stepping may clarify whether treatments that are effective for other forms of hypokinesia, such as dopaminergic medications and DBS, can also benefit parkinsonian postural instability.

## Data Availability

Data and custom software routines are available by contacting the corresponding author.

## Acknowledgments

We thank Paul Greene and Ash Rao for discussion.

## Authors’ Roles

1. Research project: A. Conception, B. Organization, C. Execution;
2. Statistical Analysis: A. Design, B. Execution, C. Review and Critique;
3. Manuscript: A. Writing of the first draft, B. Review and Critique.

RAM: 1BC, 2AB, 3AB

JCC: 1C, 2C, 3B

AW: 1C

GMM: 1BC, 3C

PM: 1ABC, 2ABC, 3B

## Financial Disclosures for Preceding 12 Months

Stock Ownership in medically-related fields: RAM: None; JCC: None; AW: None; GMM: None; PM: None

Intellectual Property Rights: RAM: None; JCC: None; AW: None; GMM: None; PM: None

Consultancies: RAM: None JCC: None; AW: None; GMM: None; PM: None

Expert Testimony: RAM: None, JCC: None; AW: None; GMM: None; PM: Yes

Advisory Boards: RAM: Data Safety Monitoring Board, Synerfuse, Inc. JCC: None; AW: None; GMM: None; PM: None

Partnerships: RAM: None JCC: None; AW: None; GMM: None; PM: None

Contracts: RAM: None JCC: None; AW: None; GMM: None; PM: None

Honoraria: RAM: None JCC: None; AW: None; GMM: None; PM: None

Royalties: RAM: None JCC: None; AW: None; GMM: None; PM: None

Grants: RAM: MnDRIVE; JCC: None; AW: None; GMM: None; PM: NINDS

Other: RAM: None JCC: None; AW: None; GMM: None; PM: None

## Financial Disclosure/Conflict of Interest

The authors do not have any financial information to disclose related to the work described in this article.

## Funding Sources

This study was supported by grants from the Parkinson’s Disease Foundation and the Doris Duke Charitable Foundation.

## Notes

### Competing Interest Statement

The authors have declared no competing interest.

### Author Declarations

The study protocol was approved by the Columbia University Medical Center Institutional Review Board.

